# Toward more realistic social distancing policies via advanced feedback control

**DOI:** 10.1101/2022.05.25.22275562

**Authors:** Cédric Join, Alberto d’Onofrio, Michel Fliess

## Abstract

A continuously time-varying transmission rate is suggested by many control-theoretic investigations on non-pharmaceutical interventions for mitigating the COVID-19 pandemic. However, such a continuously varying rate is impossible to implement in any human society. Here, we significantly extend a preliminary work (M. Fliess, C. Join, A. d’Onofrio, Feedback control of social distancing for COVID-19 via elementary formulae, MATHMOD, Vienna, 2022), based on the combination of flatness-based and model-free controls of the classic SIR model. Indeed, to take into account severe uncertainties and perturbations, we propose a feedback control where the transmission rate, *i*.*e*., the control variable, is piecewise constant. More precisely, the transmission rate remains constant during an appreciable time interval. Strict extended lockdowns may therefore be avoided. The poor knowledge of fundamental quantities such as the rate of infection hinders a precise calibration of the transmission rate. Thus, the results of our approach ought therefore not to be regarded as rules of action to follow accurately but as a guideline for a wise behavior.

## I. Introduction

The social distancing strategies and in particular the severe lockdown measures due to the worldwide COVID-19 pandemic (see, *e*.*g*., [Adolph et al.(2021)]) have stimulated a huge number of mathematically oriented investigations among which we select control-theoretic publications: See, *e*.*g*., [Al-Radhawi et al.(2022)], [Ames et al.(2020)], [Angulo et al.(2021)], [Berger(2022)], [Bisiacco & Pillonetto(2021)], [Bisiacco et al.(2022)], [Bliman & Duprez(2021)], [Bliman et al.(2021)], [Bonnans & Gianatti(2020)], [Borri et al.(2021)], [Charpentier et al.(2020)], [Dias et al.(2022)], [Di Lauro et al.(2021a)], [Di Lauro et al.(2021b)], [Efimov & Ushirobira(2021)], [Esterhuizen et al.(2021)], [Fliess et al.(2022)], [Gevertz et al.(2021)], [Greene & Sontag(2021)], [Ianni & Rossi(2021)], [Jing et al.(2021)], [Köhler et al.(2021)], [McQuade et al.(2021)], [Morato et al.(2020a)], [Morato et al.(2020b)], [Morgan et al.(2021)], [Morris et al.(2021)], [O’Sullivan et al.(2020)], [Péni et al.(2020)], [Pillonetto et al.(2021)], [Sadeghi et al.(2021)], [Sontag(2021)], [Stella et al.(2022)], [Tsay et al.(2020)]. Most diverse viewpoints have been developed. Those studies however do not seem to exert any influence on policy-makers (see, *e*.*g*., [Casella(2021)] for some explanations). Our aim is to start drawing a roadmap in order to change this state of affairs.

Like many authors in the above–mentioned works, we select the classic SIR compartmental model [Kermack & McKendrick(1927)] (see also, *e*.*g*., [Brauer & Castillo-Chavez(2012)], [Hethcote(2000)], [Murray(2002)]). An excellent justification for employing such a simple model has been presented by Sontag [Sontag(2021)]: *The social and political use of epidemic models must take into account their degree of realism. Good models do not incorporate all possible effects, but rather focus on the basic mechanisms in their simplest possible fashion. Not only it is difficult to model every detail, but the more details the more the likelihood of making the model sensitive to parameters and assumptions, and the more difficult it is to understand and interpret the model as well as to play what-if scenarios to compare alternative containment policies. It turns out that even simple models help pose important questions about the underlying mechanisms of infection spread and possible means of control of an epidemic*. In addition, the rate of infection and other fundamental quantities are difficult, if not impossible, to know precisely (see, *e*.*g*., [Havers et al.(2020)], [Pérez-Rechel et al.(2021)], [Perkins et al.(2020)]). This epistemological hindrance to mathematical epidemiology provides further legitimacy for using a parsimonious modeling.

The *transmission rate β*, which corresponds to the social interactions and to infection probability per contact, is chosen as the control variable, like in most papers which are quoted above. Thus, our work can be framed in the field of behavioral epidemiology of infectious disease (see, *e*.*g*., [Manfredi & d’Onofrio(2013)]).

The SIR model happens then to be (*differentially) flat*. This concept [Fliess et al.(1995)], [Fliess et al.(1999)] (see also the books [Sira-Ramírez & Agrawal(2004)], [Lévine(2009)], [Rigatos(2015)], [Rudolph(2021)]) has given rise, as is commonly known, to numerous concrete applications mainly in engineering (see, *e*.*g*., [Bonnabel & Clayes(2020)] for a recent excellent publication about cranes), but also in other domains (see, *e*.*g*., [Guéry-Odelin et al.(2019)] in quantum physics). Also of particular interest here is its use [Hametner et al.(2022)] for COVID-19 predictions.

Take a flat system with a single control variable *u*(*t*) and a flat output variable *y*(*t*). From a suitable reference trajectory *y*^*\**^(*t*), *i*.*e*., a suitable time-function, the corresponding open-loop nominal control variable *u*^*\**^(*t*) is derived at once from the flatness property. Severe uncertainties, like model mismatch, poorly known initial conditions, external disturbances, …, prompt us to mimic what has been already done by [Villagra & Herrero-Pérez(2012)], [Fliess et al.(2021)], [Fliess et al.(2022)], *i*.*e*., to close the loop via *model-free* control in the sense of [Fliess & Join(2013)], [Fliess & Join(2021)]. Among the numerous remarkable concrete applications of this approach let us cite two recent ones in different domains (see, *e*.*g*., [Kuruganti et al.(2021)], [Lv et al.(2022)], [Sancak et al.(2021)], [Wang et al.(2021)]), and, especially here, mask ventilators for COVID-19 patients [Truong *et al*.(2021)]. Take another output variable *z*(*t*) and its corresponding reference trajectory *z*^*\**^(*t*). The feedback loop, which relates Δ*β* = *β − β*^*\**^ and the tracking error Δ*z* = *z − z*^*\**^, is expressed as an *intelligent Proportional*, or *iP*, controller [Fliess & Join(2013)]. This is much easier to implement than traditional PI and PID controllers (see, *e*.*g*., [Åström & Murray(2008)]) and ensures local stability around *z*^*\**^ with a remarkable level of robustness. Inspired by techniques in [Lafont et al.(2015)] and [Join et al.(2022)], which were performed in practice for a greenhouse and ramp metering on highways, we close the loop such that the control variable *u* = *u*^*\**^ + Δ*u*

- takes only a finite number of numerical values,
- remains constant during some time interval.^1^

These features, which are new to the best of our knowledge, imply a limited number of different non-pharmaceutical interventions which moreover are not too severe. They might therefore be socially acceptable. Only low computing capacity is necessary for conducting numerous *in silico* experiments.^2^

Our paper is organized as follows. The flatness property of the SIR model is shown in Section II. An open-loop strategy is easily derived in Section II-B. Section III introduces closed-loop control via model-free control. Several computer simulations are displayed in Section IV.^3^ Section V is devoted to a discussion of the possible implications of our approach.

## II. SIR and open-loop control

### A. Flatness

The well known SIR model,^4^ which studies the populations of *susceptible*, whose fraction is denoted as *S, infectious*, whose fraction is denoted as *I*, and *recovered* or *removed*, whose fraction is denoted as *R*), reads:

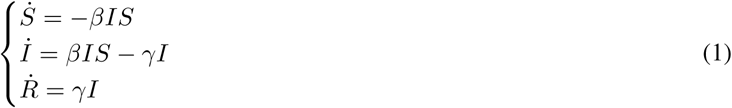

The transmission rate *β* and the *recovery/removal rate γ* are positive. Equation (1) yields that *S* + *I* + *R* is constant. We may set

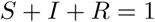

Straightforward calculations yield:

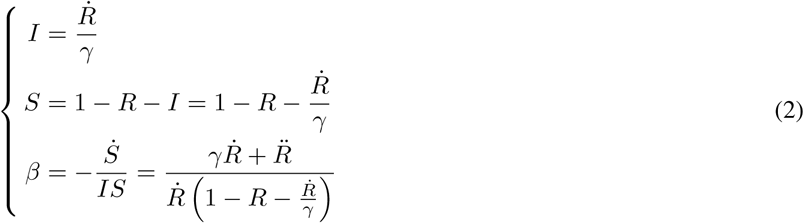

The system variables *I, S* and *β* may be expressed as rational *differential* functions of *R, i*.*e*., as rational functions of *R* and its derivatives up to some finite order. In other words, System (1) is, as already observed [Fliess et al.(2022)], flat, and *R* is a flat output.

*Remark 1:* The SEIR^5^ model (see, *e*.*g*., [Brauer & Castillo-Chavez(2012)], [Hethcote(2000)]) is a rather popular extension of the SIR model:

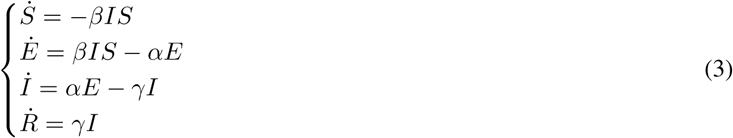

where *α >* 0 is an additional parameter. Now

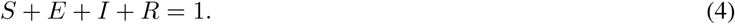

Equations (3)-(4) show that the SEIR model is flat and that *R* is again a flat output:

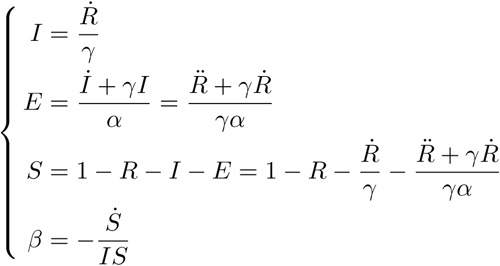

### B. Elementary formulae for open-loop control

Select the following decreasing exponential reference trajectory,^6^ where *λ >* 0,

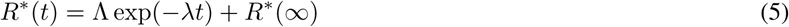

The quantities Λ, *R*^*\**^(*∞*) = lim_*t→*+*∞*_ *R*^*\**^(*t*) are given below. Formulae (2) and (5) yield

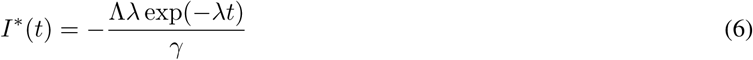

Formula (6) implied of course that Λ *<* 0. The fundamental constraint

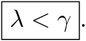

follows at once from the differential inequality *İ > −γI*. Moreover

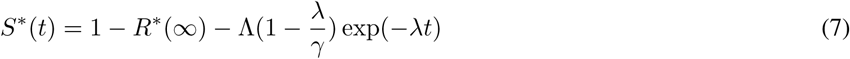

The corresponding open loop control reads

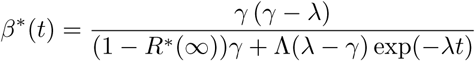

Note that *β*^*\**^(*t*) *>* 0. Thus

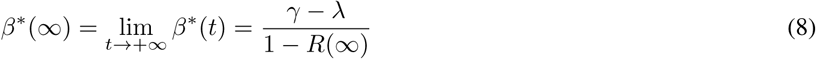

The parameter Λ in Equation (5) may be expressed thanks to Equation (6):

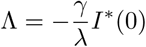

Thus

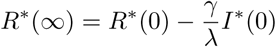

In order to avoid discontinuities at time *t* = 0, set *R*^*\**^(0) = *R*(0), *I*^*\**^(0) = *I*(0). The following expressions for *λ* and *R*^*\**^(*∞*) will be used:

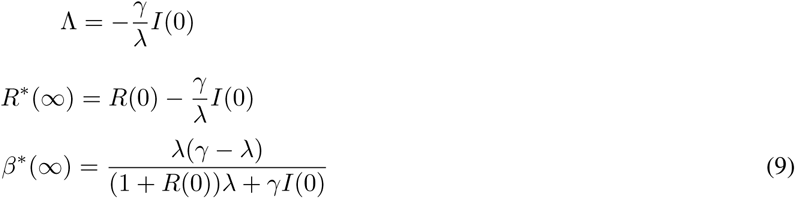

Introduce the more or less precise quantity *β*_accept_: It is the “harshest” social distancing protocols which is “acceptable” in the long run. Equation (9) yields

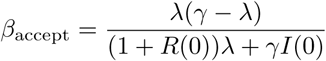

and an algebraic equation of degree 2 for determining *λ*

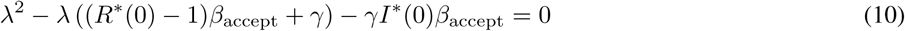

The two roots of Equation (10) are real. The only positive one

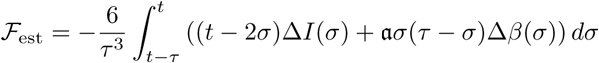

is the value of *λ* we are looking for. The corresponding reference trajectory and nominal open-loop control follow at once.

## III. Closed-loop control

In order to take into account unavoidable mismatches and disturbances, introduce the *ultra-local* model [Fliess & Join(2013)]

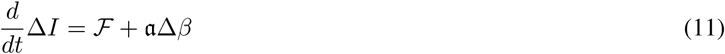

where

- Δ*I* = *I − I*^*\**^, Δ*β* = *β − β*^*\**^,
- the constant parameter *a*, which does not need to be precisely determined, is chosen such that the three terms in Equation (11) are of the same magnitude.
- *ℱ* subsumes the unknown internal structure and the external disturbances.
- An estimate *ℱ*_est_ of *ℱ* is given [Fliess & Join(2013)] by the integral

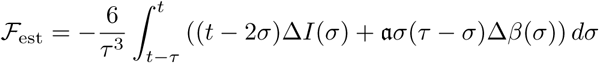

which in practice may be computed via a digital filter.

An *intelligent proportional*, or *iP*, controller [Fliess & Join(2013)] reads

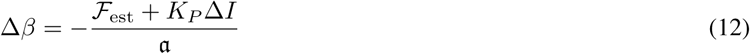

where *K*_*P*_ a classic tuning gain and *ℱ*_est_ an estimate of *ℱ*. Combining Equations (11) and (12) yields

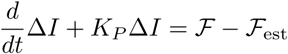

If *K*_*P*_ *<* 0, and if the estimate *ℱ*_est_ is “good”, *i*.*e*., *ℱ*_est_ *≈ ℱ*, then

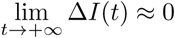

Thus local stability around 0 is ensured in spite of mismatches and external disturbances.

## IV. Computer simulations^7^

Set in Equation (1) 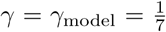. The rate *I*(*t*) of infected people is assumed to be counted every 2 hours.^8^ The iP (12) is employed in all the scenarios below, with *a* = 0.01 and *K*_*P*_ = 15*a*.

### A. Unrealistic scenarios

A naïve application of Section II-B leads to a continuous evolution of the control variable *β, i*.*e*., of the social distancing. This is obviously impossible to implement in real life.

#### 1) Scenario 1

Let us first assume that *I*(0) and *R*(0) are perfectly known. This initial time 0 is set after 35 or 45 days of epidemic spreading, where *β* = 3.6*γ*_model_. Thus *I*(0) after 35 days is less than after 45 days. Figures 1 and 2 display excellent results, where *β*_acccept_ = 0.95*γ*_model_. Note that, even here, closed-loop control is necessary in order to counteract the unavoidable rounding errors.

**Fig. 1:**
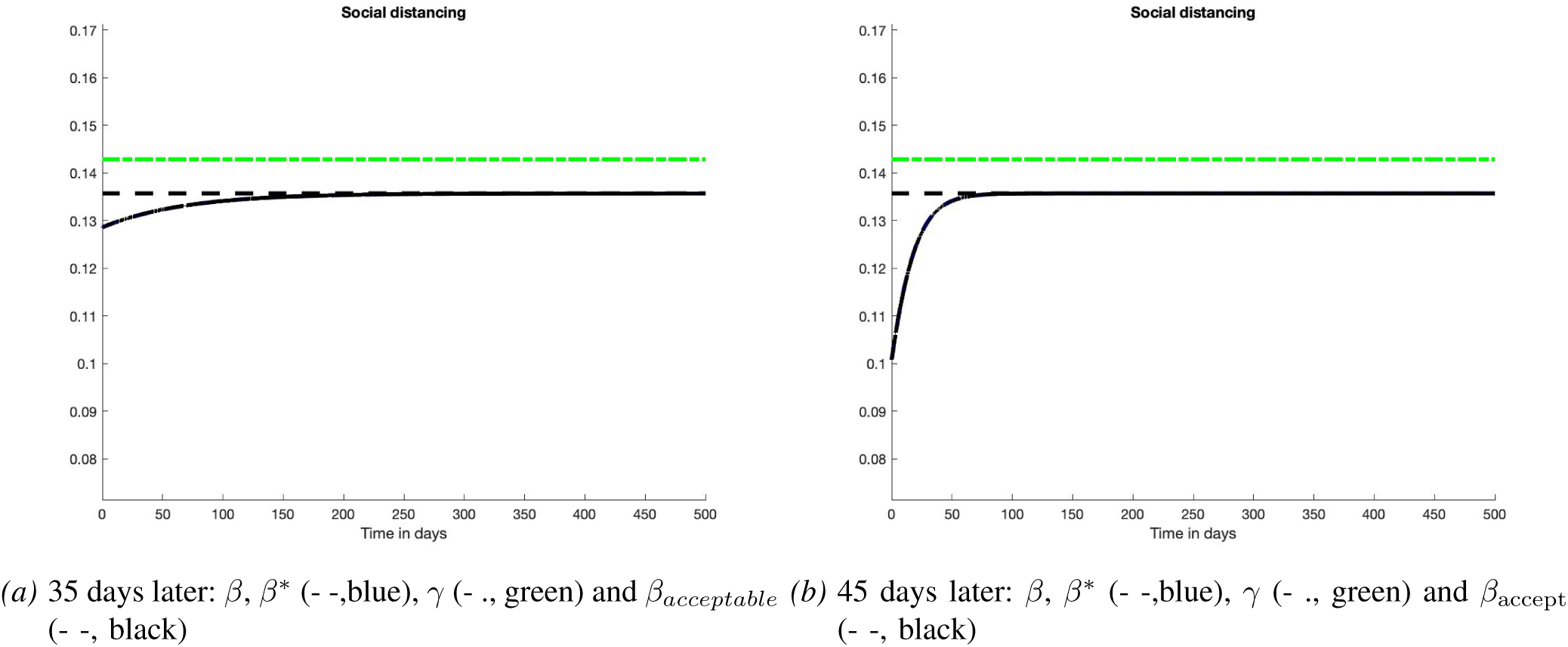
Scenario 1: Open-loop control

**Fig. 2:**
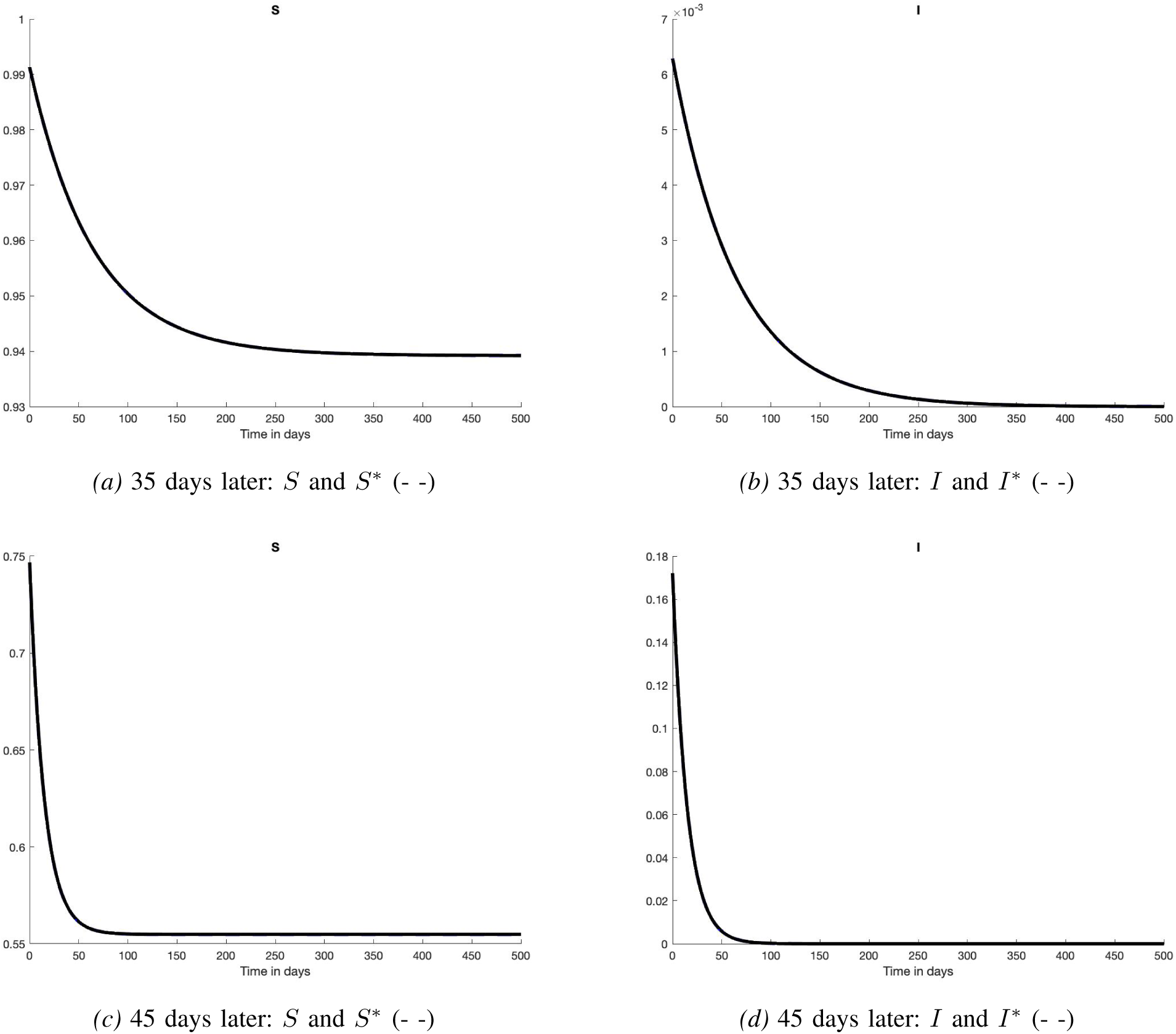
Scenario 1: States

### B. Scenario 2

The initial time is set after 35 days of epidemic spreading. Introduce some mismatches:

- *γ*_real_ = 1.05*γ*_model_ (Figures 3-(a) and 4-(a)-(b)),
- *γ*_real_ = 0.95*γ*_model_ (Figures 3-(b) and 4-(c)-(d)).

It is obvious then that the tracking of *I*^*\**^ yields to deviate from *R*^*\**^ and 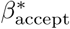.

**Fig. 3:**
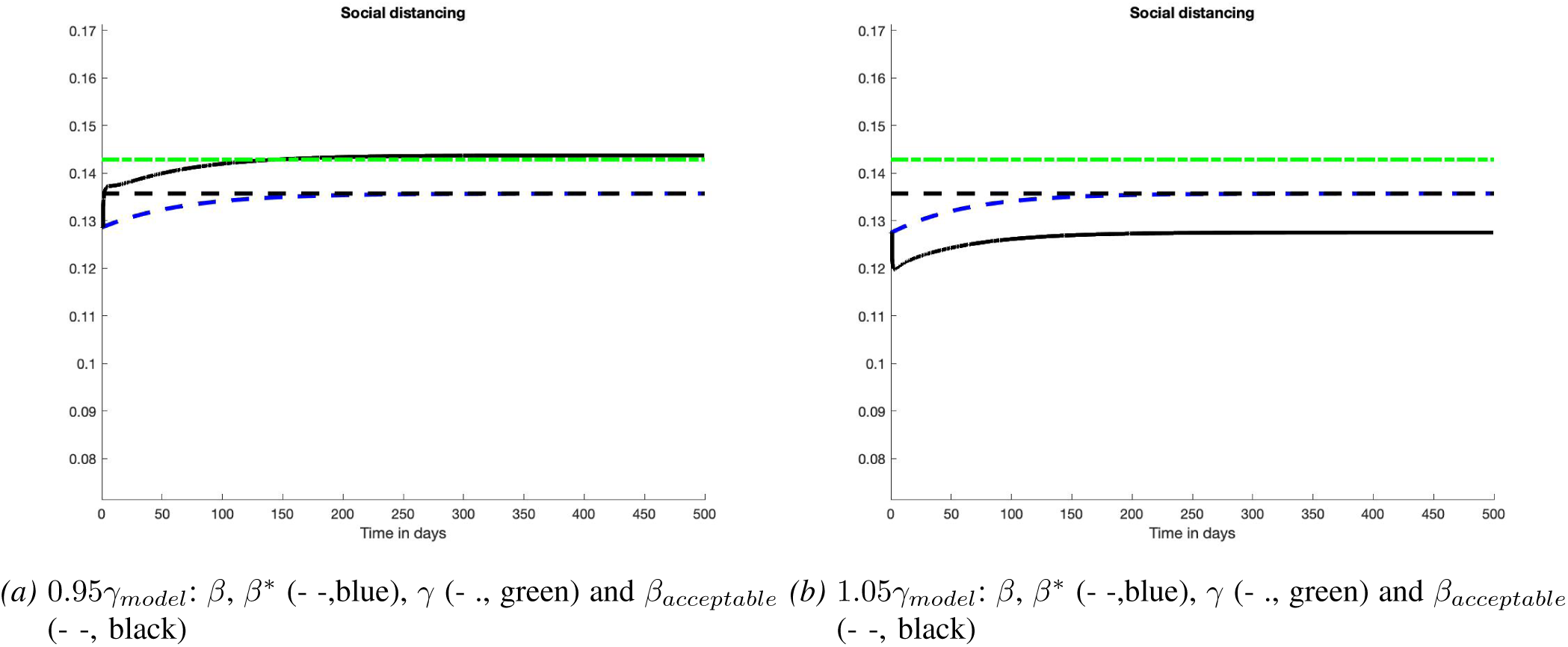
Scenario 2: Control

**Fig. 4:**
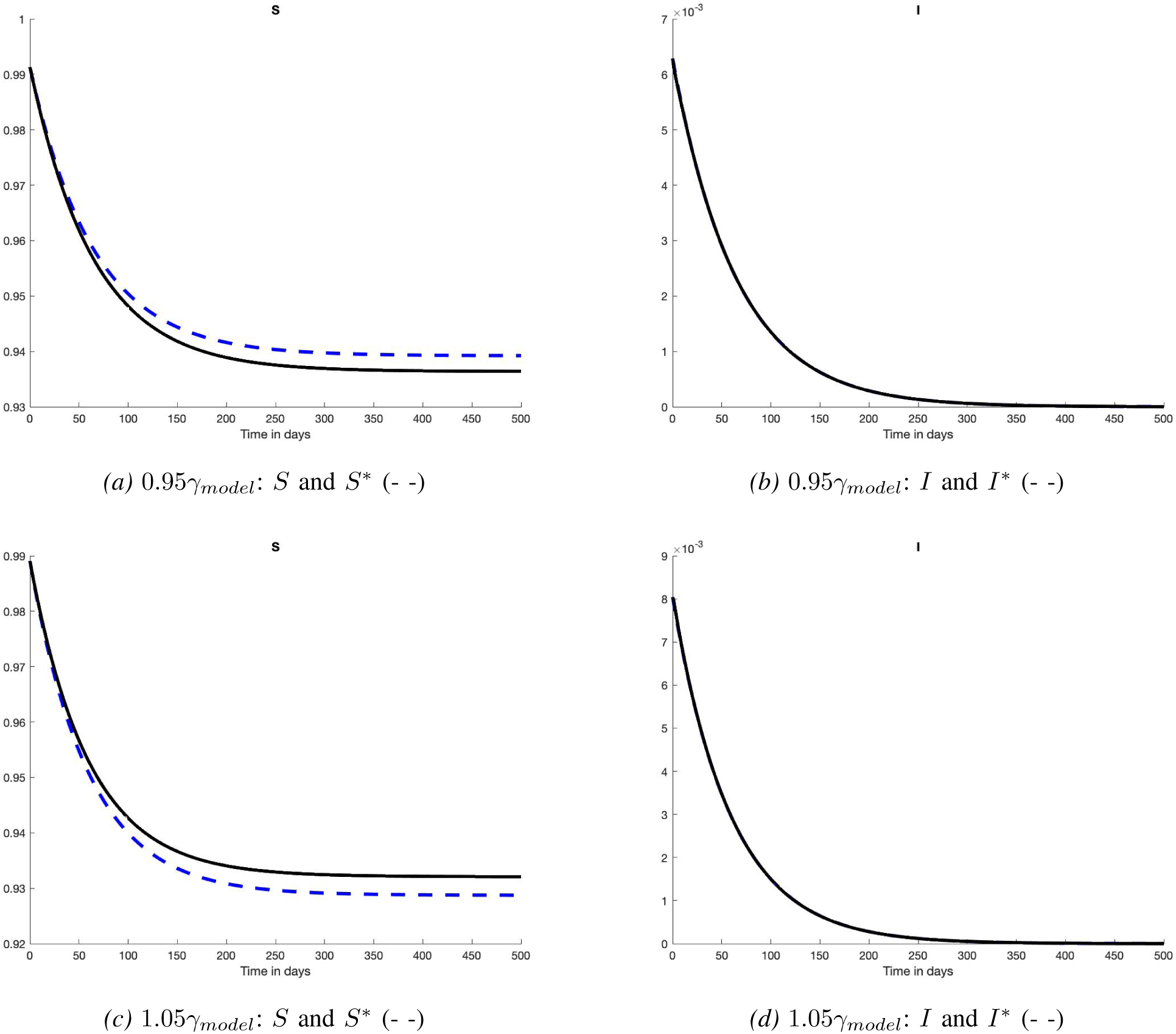
Scenario 2: States

### C. Less unrealistic scenarios

Take the same conditions as in Section IV-A.1: no model mismatch, perfectly known initial conditions, the initial time 0 is set after 35 or 45 days of epidemic spreading.

1) *Scenario 3:* Allow only a finite number of numerical values of the control variable *β*. The control variable *β* takes 50 uniformly distributed values between 0.5*γ*_model_ and 1.2*γ*_model_. Figures 5 - 6 (35 days) and 7 - 8 (45 days) display excellent results.
2) *Scenario 4:* Allow *β* now to take any value between 0.5*γ*_model_ and 1.2*γ*_model_ but to change only every 14 days. Figures 9 - 10 (35 days) and Figures 11 - 12 (45 days) display rather violent alternations in the social distancing rules. The tracking of *I*^*\**^ remains good.

**Fig. 5:**
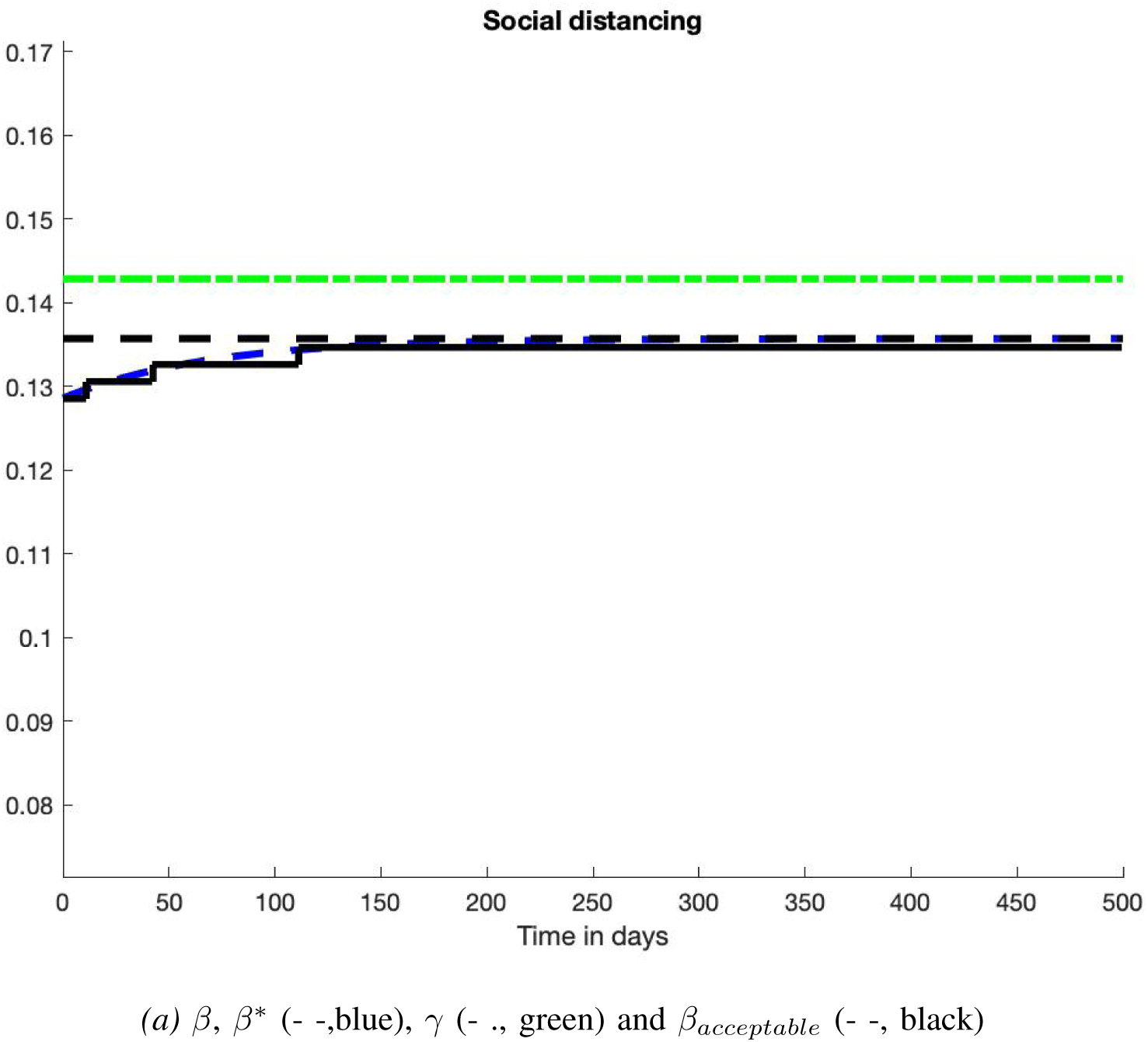
Scenario 3: Control

**Fig. 6:**
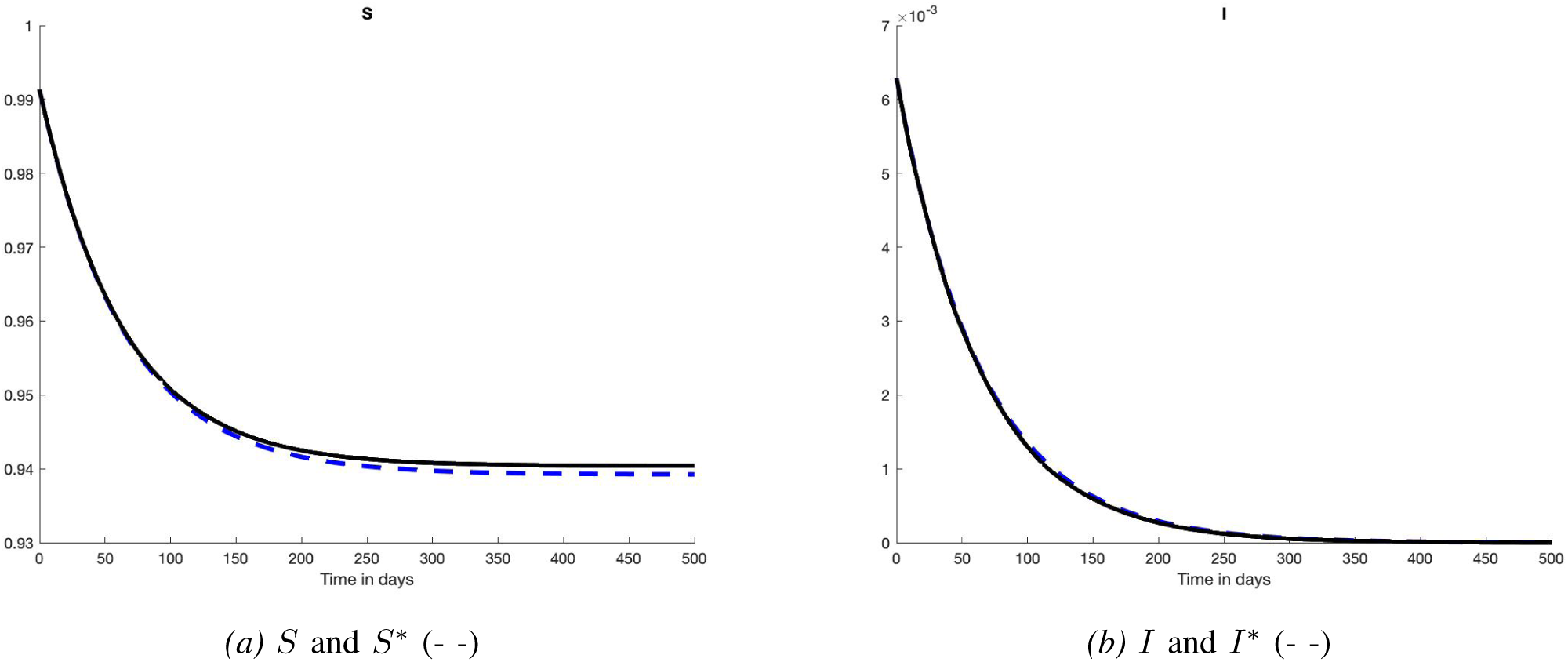
Scenario 3: States

**Fig. 7:**
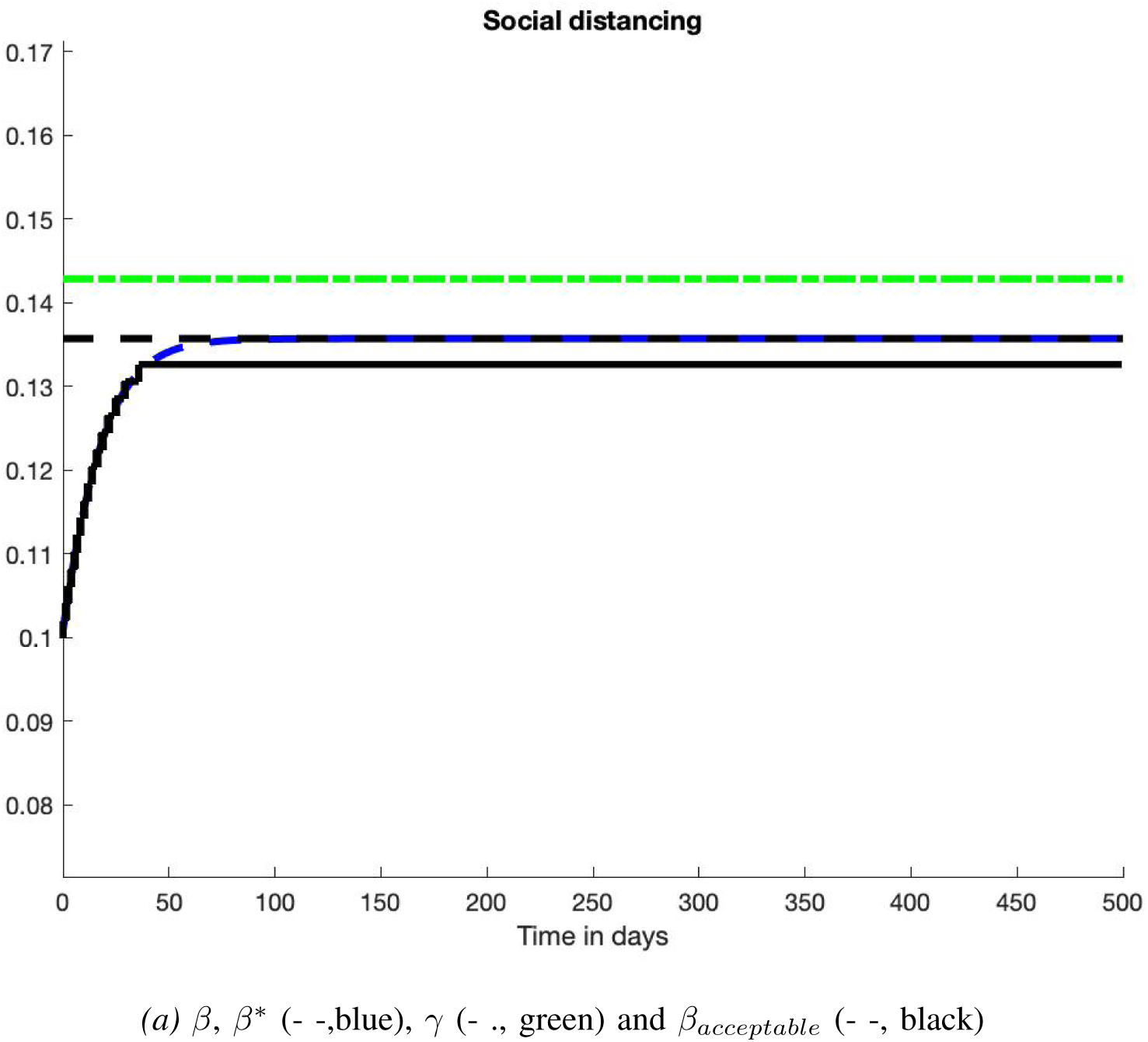
Scenario 4: Control

**Fig. 8:**
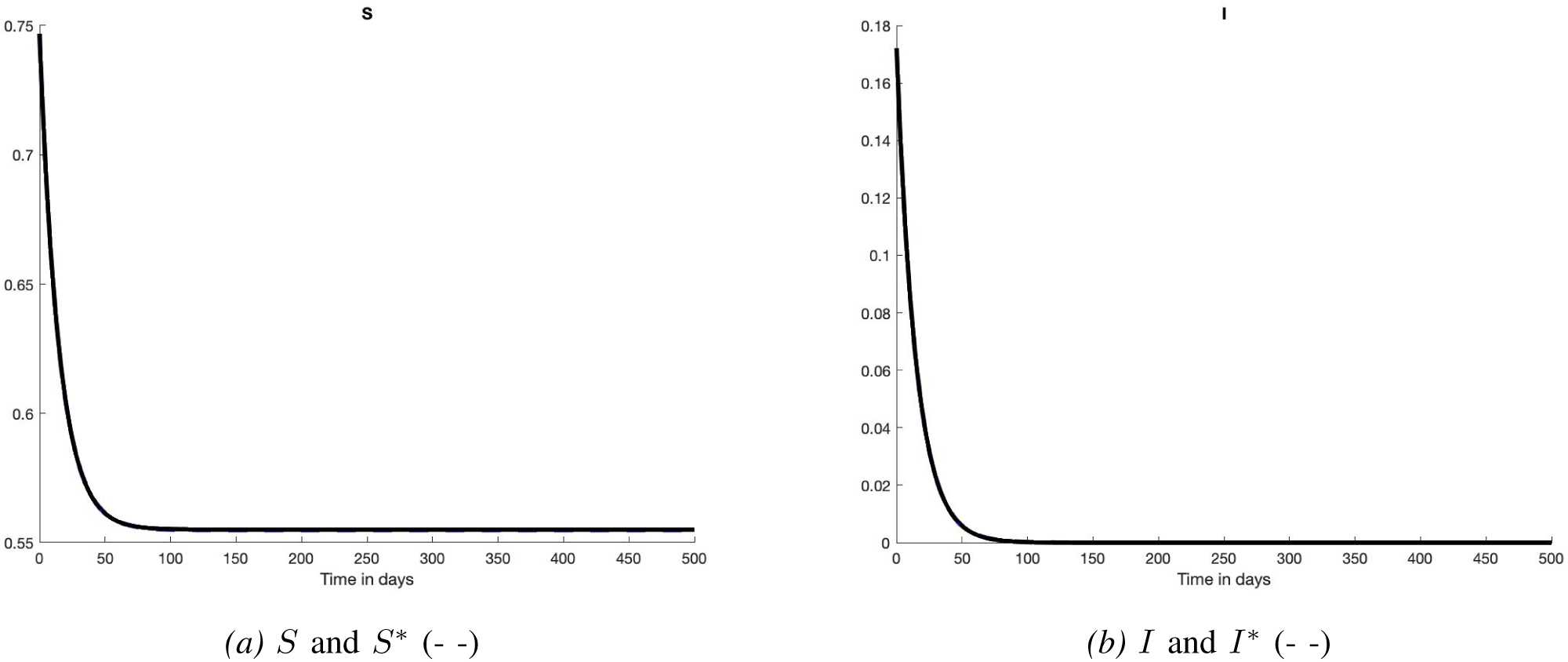
Scenario 4: States

**Fig. 9:**
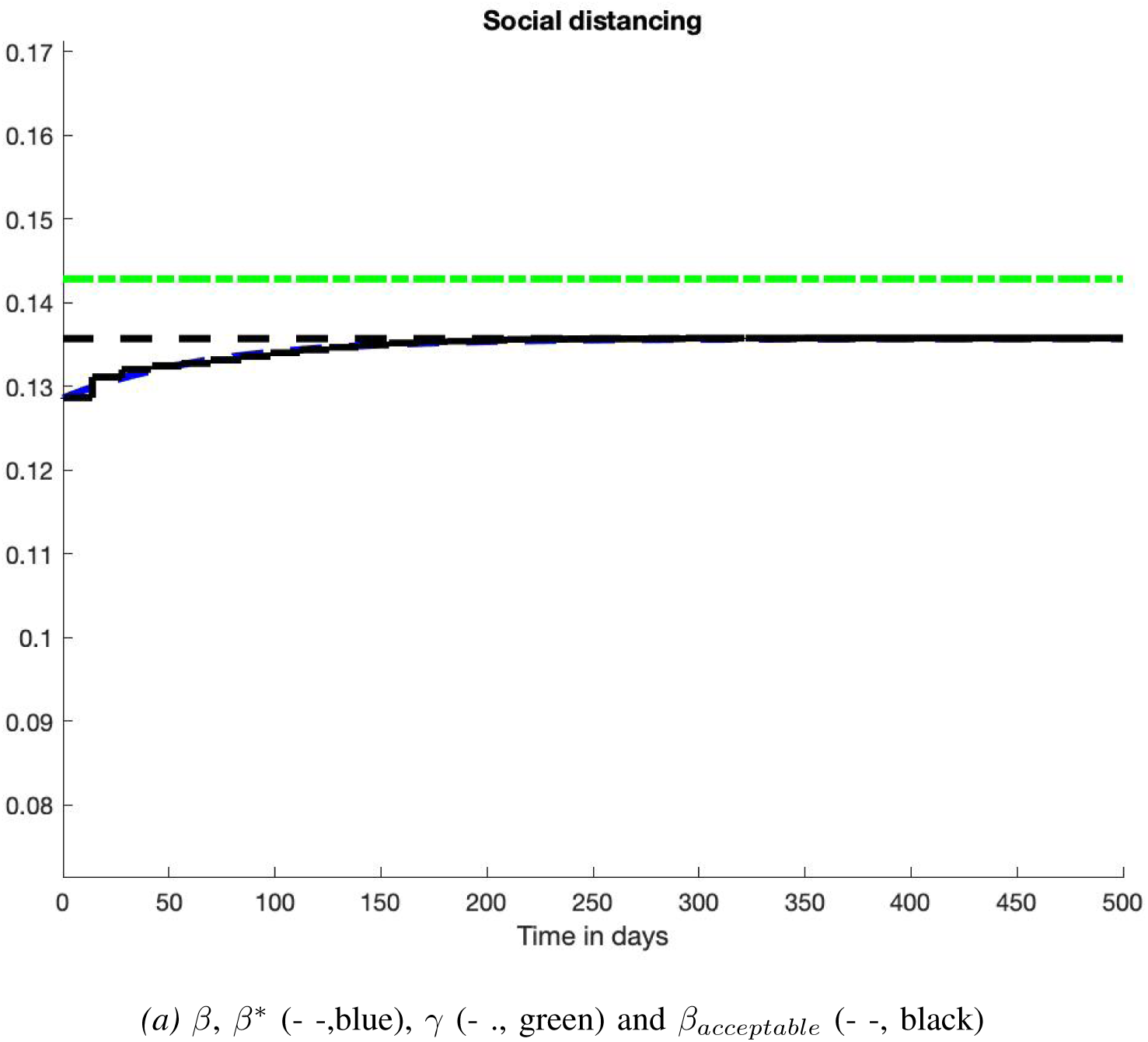
Scenario 5: Control

**Fig. 10:**
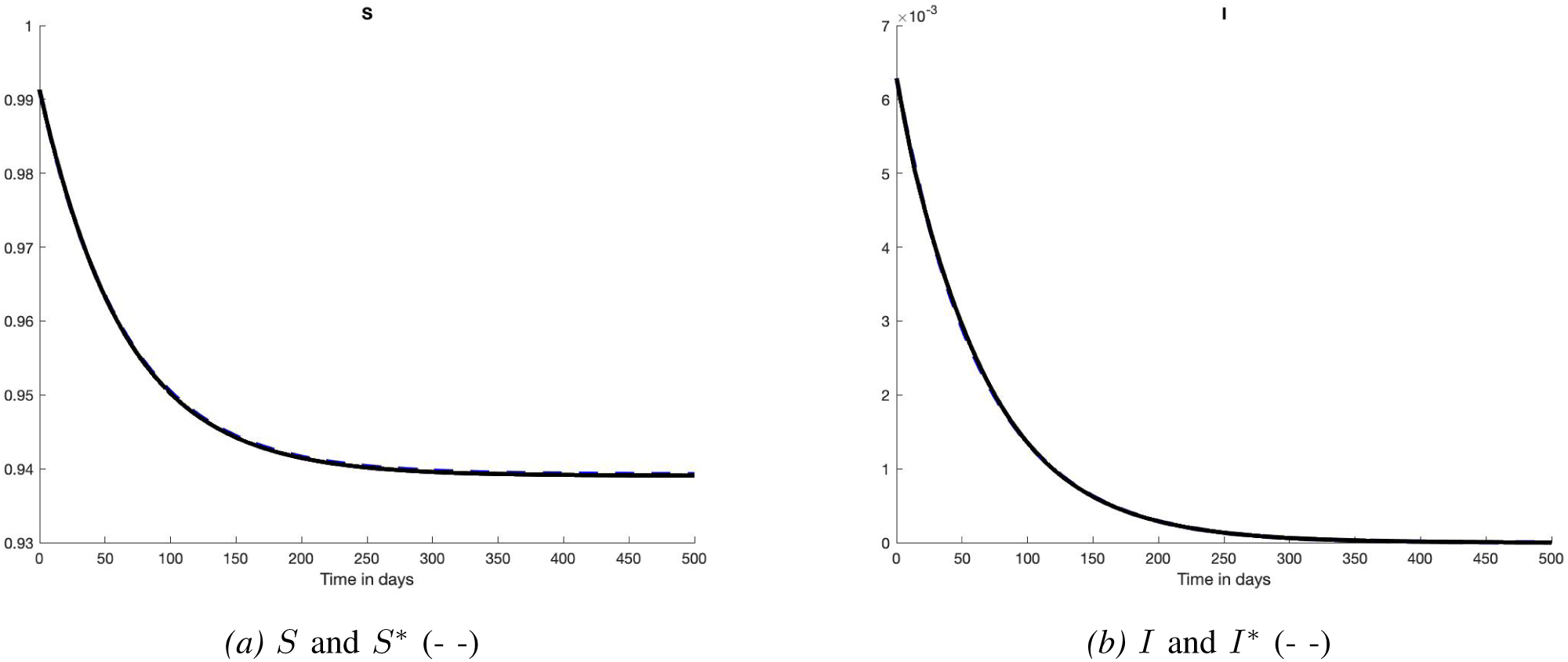
Scenario 5: States

**Fig. 11:**
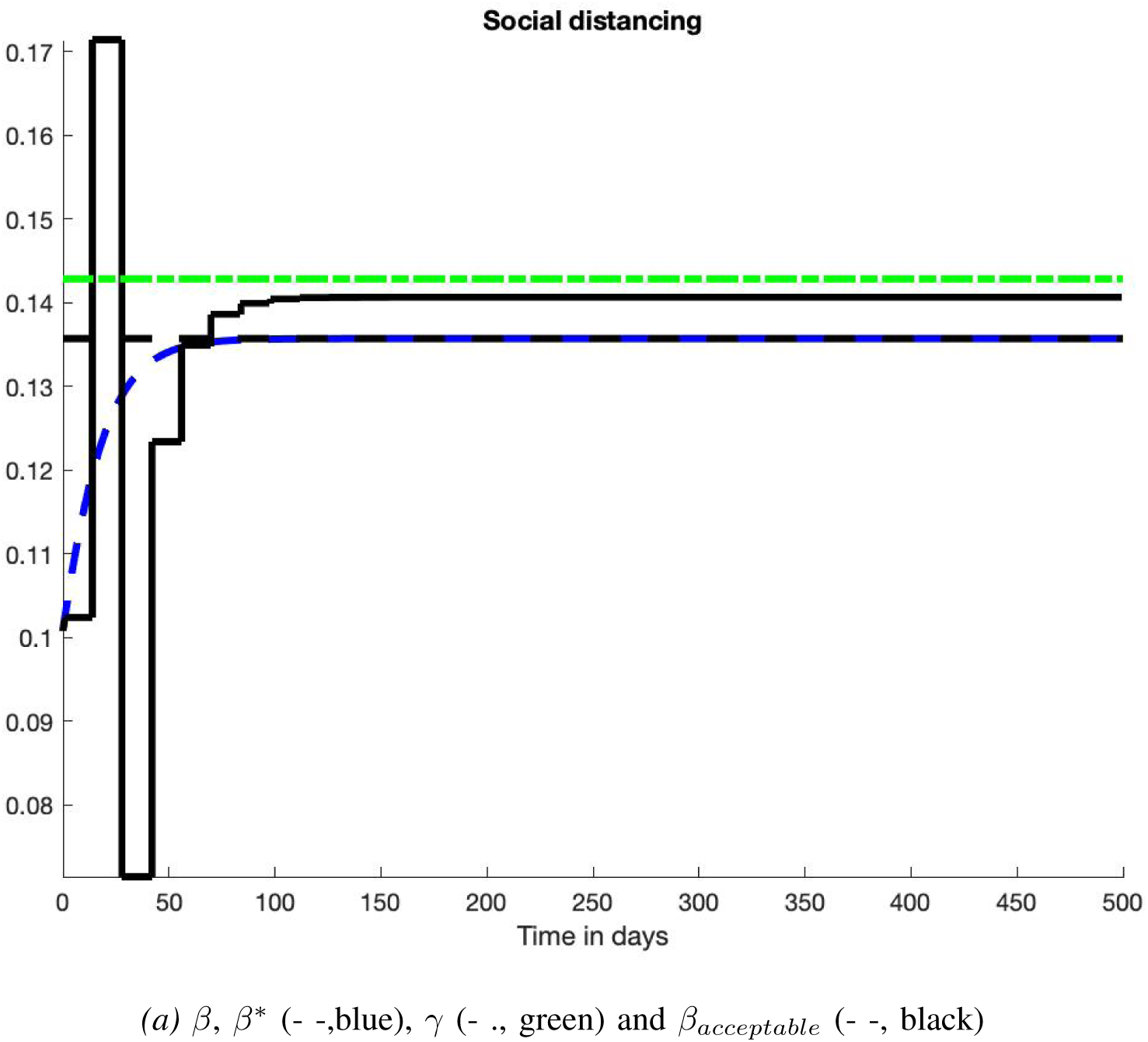
Scenario 5: Control

**Fig. 12:**
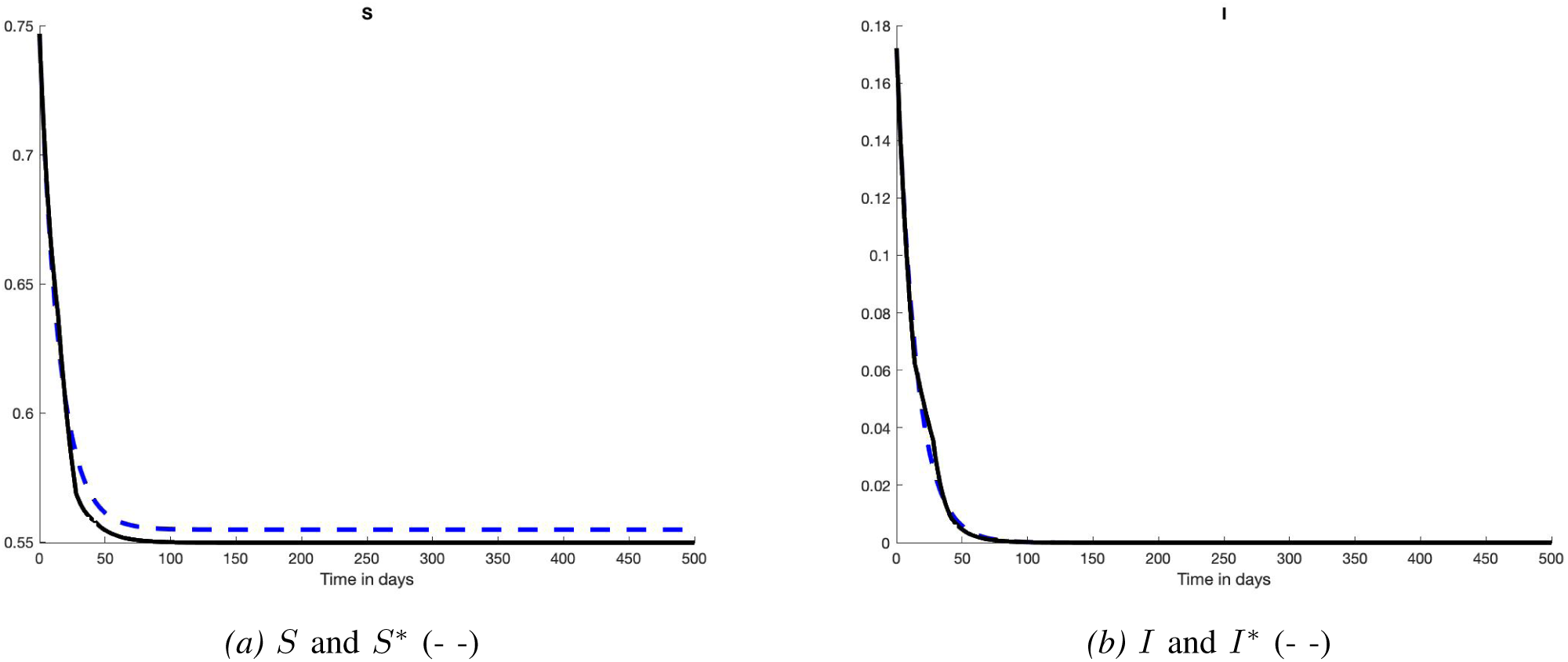
Scenario 5: States

### D. Scenarios 5-6: A more realistic policy

We follow Section IV-C and combine the conditions on the control variable *β* of Sections IV-C.1 and IV-C.2. Thus *β* takes only a finite number of values and change every 14 days. Correct results are provided in Figures 13 - 14 (35 days) and Figures 15 - 16 (45 days). The tracking of *I*^*\**^ is still excellent.

**Fig. 13:**
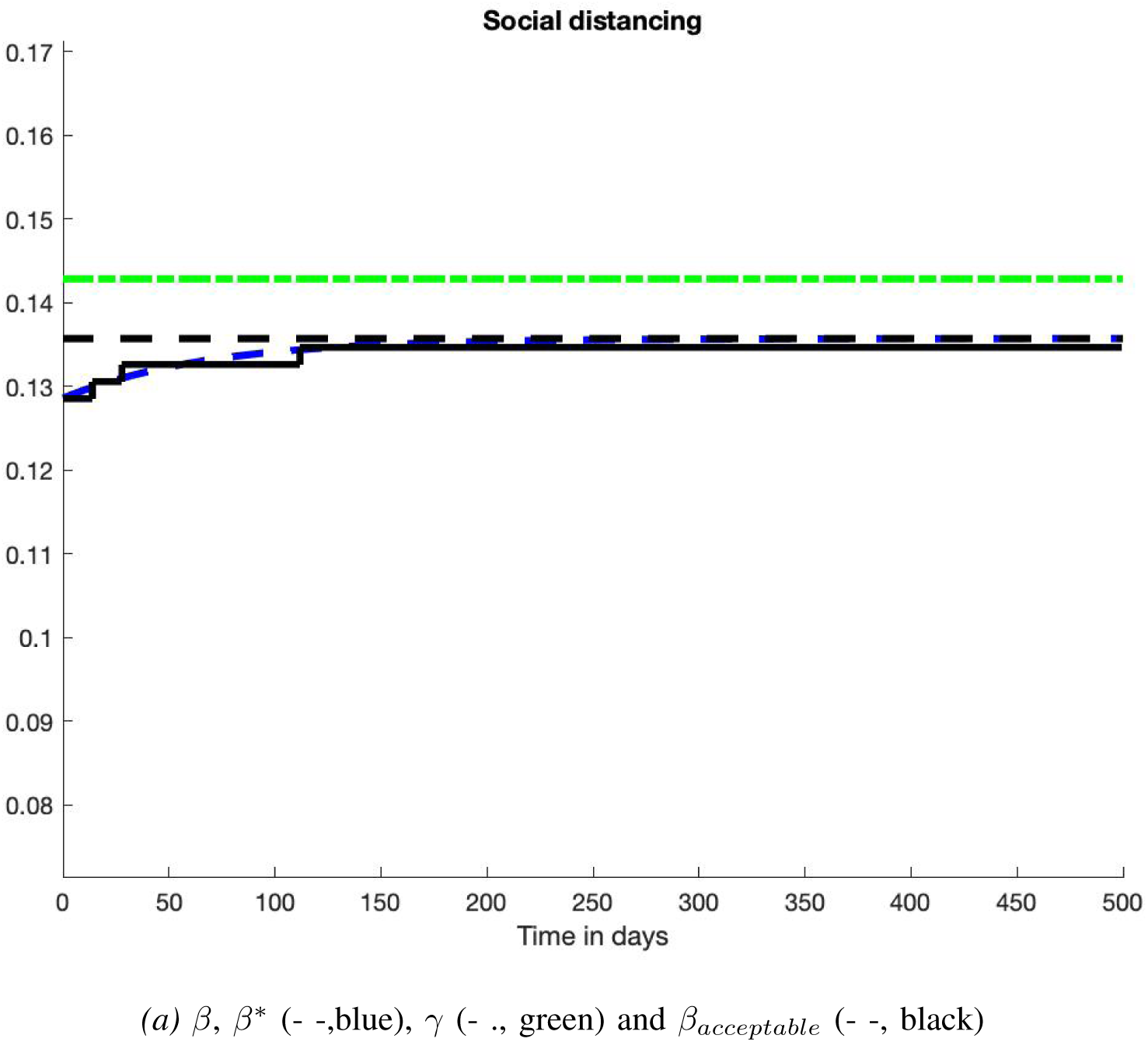
Scenario 6: Control

**Fig. 14:**
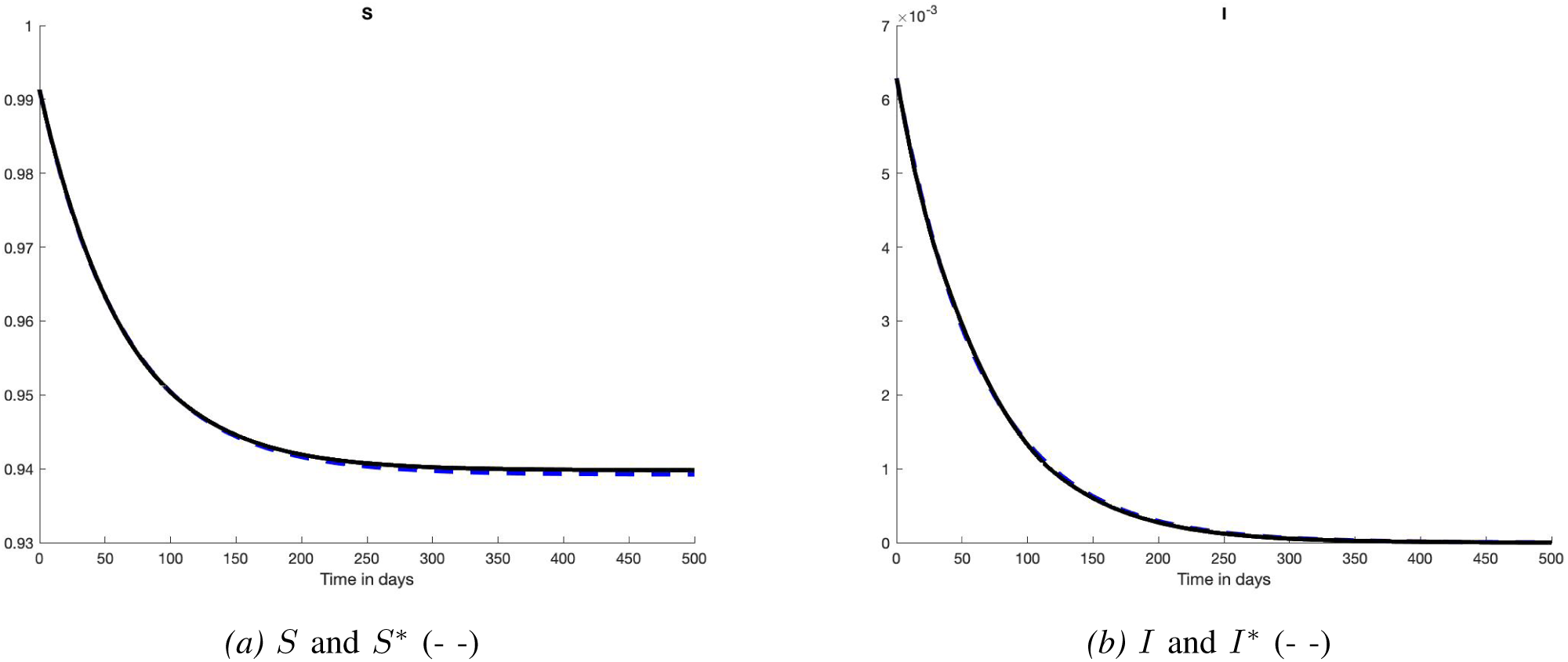
Scenario 6: States

**Fig. 15:**
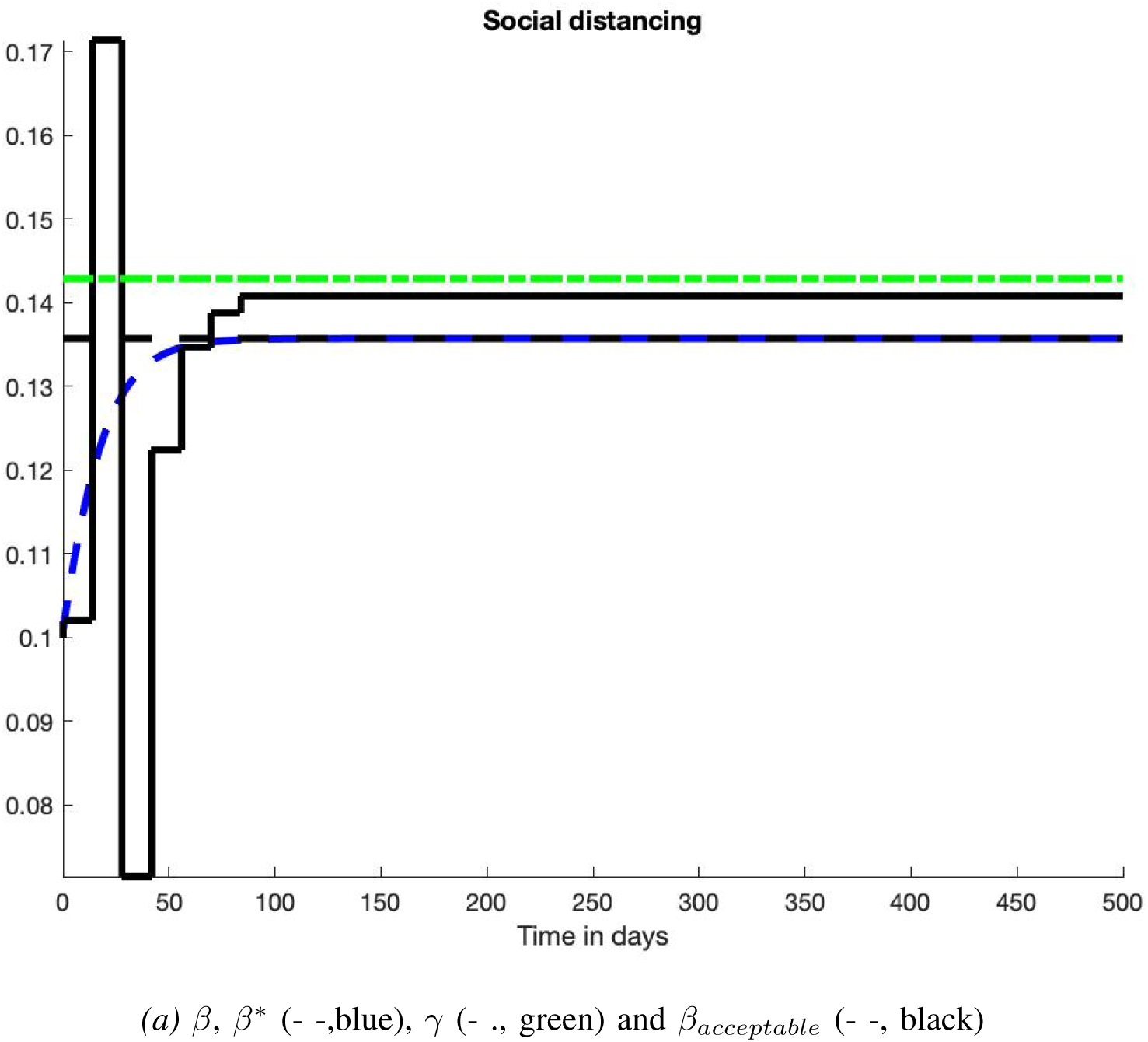
Scenario 6: Control

**Fig. 16:**
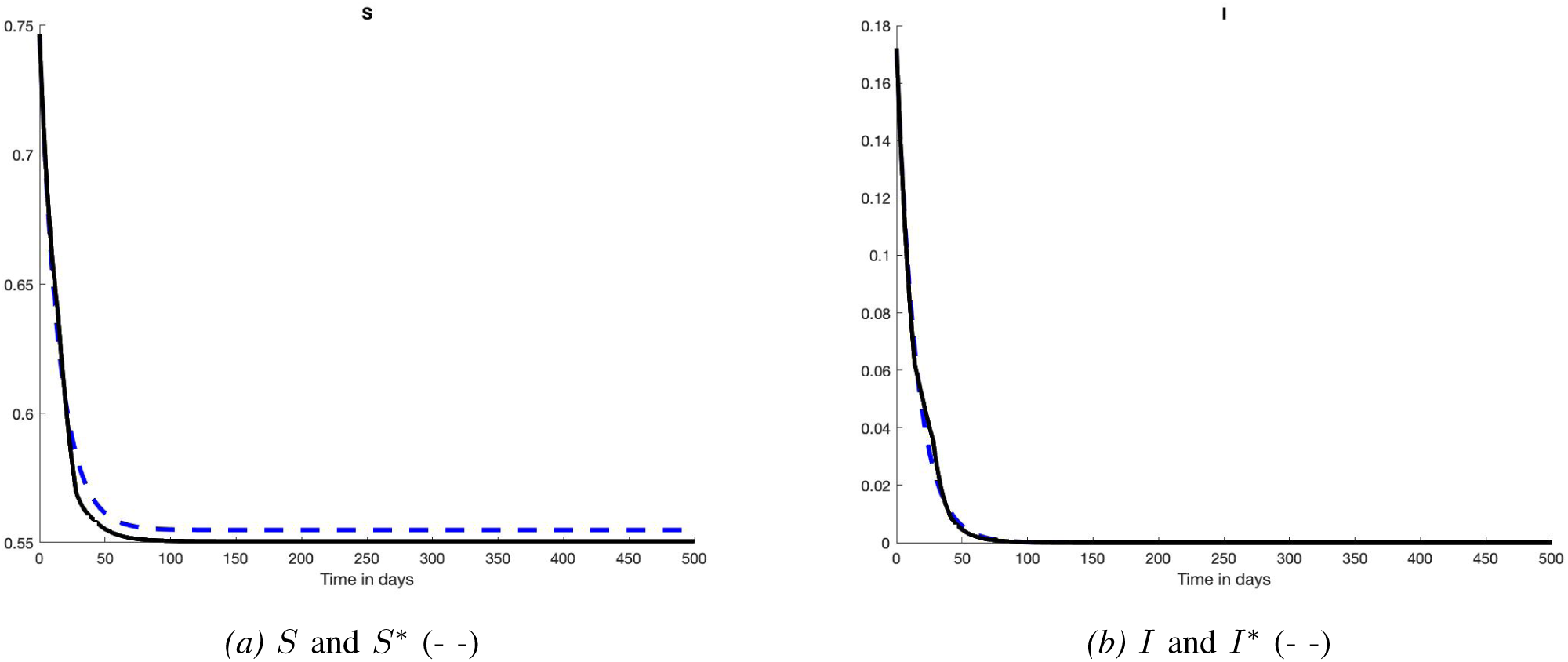
Scenario 6: States

## V. Conclusion

We have designed a disease mitigation strategy, which is based on recent advances in mathematical control, where

- continuous manipulations of non-pharmaceutical interventions are avoided,
- severe and long lockdowns are replaced by more subtle alternations of more or less strict social distancing measures.

Today empirical control strategies are adopted in practice. However, they are based an the principle of trial and error, which is very risky in epidemic context. Thus, the introduction of more rigorous but realistically constrained approaches might be of interest for policy–makers. Another point to be stressed is that our overall results are also robust in presence of uncertainties in key parameters.

Several other questions arise:

- How should one modify the above approach and simulations when vaccinations and variants are taken into account? See, *e*.*g*., [Arruda et al.(2021)], [Kopfovál et al.(2021)], [Laguzet & Turinici(2015)], [Moore et al.(2021)], [d’Onofrio et al.(2007)], [Ramos et al.(2021)] for some preliminary modeling issues.
- Another interrogation is about the interpretation of the numerical values of the control variable *β*. What is, for instance, the influence of closing nightclubs as done in France and elsewhere? Available estimation techniques would suffer from the poor knowledge of *I*, and therefore of *R* and *S*.^10^

## Data Availability

All the data are available in the paper

## Footnotes

1 Two weeks in our computer simulations.

2 Computer experiments.

3 They considerably improve [Fliess et al.(2022)].

4 The notation 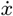 below, in Equation (1), stands for the time derivative of the time function *x*(*t*). The notation 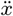 in Equation (2) designates the second order deribative.

5 *Susceptible-Exposed-Infected-Recovered/Removed*

6 Contrarily to [Fliess et al.(2022)], we do not start with *R*^*\**^ (0) = 0.

7 Contrarily to [Fliess et al.(2022)], the scenarios below do not necessarily start at the beginning of the epidemic.

8 This time lapse is of course too short in practice. It has been chosen in order to have enough points for our numerical analysis.

9 As a consequence, this is an obstacle to the implementation of the vast majority of theoretical control strategies in epidemiology, even for the remarkably different problem of vaccination awareness campaigns (see [Della Marca & d’Onofrio(2021)]).

10 This inefficiency includes the techniques employed in [Fliess et al.(2022)] where the assumed knowledge of *I* is unrealistic.

## Notes

### Competing Interest Statement

The authors have declared no competing interest.

### Funding Statement

This study did not receive any funding

